# Clinical profile and plasma Vitamin B6 levels in children with Tourette Syndrome

**DOI:** 10.64898/2026.01.14.26343936

**Authors:** Anusree A Kumar, Aisha Shaju, Varsha Vidyadharan, D Dhanasooraj, Rajith K Ravindren

## Abstract

**Background:** Tourette syndrome is a childhood-onset neuropsychiatric disorder characterised by recurrent motor and vocal tics. It shows a marked male predominance and is frequently associated with comorbid conditions such as attention-deficit/hyperactivity disorder (ADHD) and obsessive– compulsive disorder (OCD). Histaminergic dysregulation in the brain has been proposed as one of the mechanisms underlying Tourette syndrome. Vitamin B6, a key cofactor in histamine metabolism, may therefore play a contributory role in its pathophysiology.

**Method:** The clinical features of 25 children diagnosed with Tourette syndrome were assessed using the Yale Global Tic Severity Scale. Plasma vitamin B6 levels were measured using enzyme-linked immunosorbent assay (ELISA) and compared with those of a control group.

**Result:** Most participants were males, and 16% had comorbid ADHD or OCD. The most common motor tics were eye blinking, shoulder shrugging, head jerking, and orofacial movements. Frequent vocal tics included throat clearing, sniffing, uttering syllables, and breathing-related tics. Coprolalia was observed in four children. The median plasma vitamin B6 level in the Tourette syndrome group was 25.01ng/ml, which was significantly lower than the 36.33ng/ml in the control group (Mann–Whitney U = 225, p = 0.03). The rank-biserial correlation indicated a moderate effect size (r = 0.35).

**Conclusion:** Tourette syndrome in children predominantly affects males and is commonly associated with ADHD and OCD. Coprolalia-a clinically distressing symptom - was present only in a small subgroup. The lower plasma vitamin B6 levels observed in children with Tourette syndrome suggest a possible role for vitamin B6 in disease pathogenesis, potentially through its involvement in histaminergic and GABAergic neurotransmission, as well as in the modulation of neuroinflammatory processes.

## INTRODUCTION

Tourette syndrome (TS) is characterized by multiple brief, stereotypical nonrhythmic movements and vocalizations called tics that last at least a year. The prevalence is around 0.3-1% worldwide.^1^ It is seen more in males with ADHD, OCD, learning difficulties, and disruptive behavior, being the common comorbidities. Common motor tics include eye blinking, facial grimacing, nose twitching, and movements of the jaw, neck, and shoulders. Motor tics usually appear between the ages of 4 and 6 years, several years before the onset of vocal tics, which usually begin between 8 and 12 years. With increasing age, motor tics may evolve into more elaborate movements (e.g., purposeful movements or lewd gestures), and vocal tics often develop into repeating words (echolalia), phrases (palilalia), or obscene words (coprolalia). Tics have a waxing and waning character with 50 % of children achieving remission by 15 years.^2^

TS occurs due to excessive dopamine in the basal ganglia and frontostriatal pathways.^2^ Increased activity in the striatum and heightened compensatory activity of the left prefrontal cortex and subthalamic nucleus have been observed in patients with TS.^3^ Postmortem analysis of the striatal transcriptome showed increased inflammatory response in microglia. TS is normally treated by using dopamine blockers like risperidone, haloperidol, or pimozide.^2^

In addition to dopamine, histamine dysregulation has been implicated in the pathophysiology of TS.^5^ In a family with a high incidence of TS, a nonsense mutation in the histidine decarboxylase gene was identified (HDC W317X).^6^ The HDC gene is essential to convert histidine to histamine. Reduced histamine causes dysregulation of dopamine activity, thereby contributing to the development of TS.^7^ The HDC W317X mutation is rare and has not been identified outside of the index family. Other disruptions in the histamine pathway that reduce histamine may prove to be causative in the TS pathophysiology. We hypothesize that vitamin B6-through its role as a cofactor of HDC enzyme – plays a role in histamine regulation and thereby TS. This hypothesis has a potential translational significance in the management of TS.

Though the antipsychotics improve the symptoms of TS, their side effects limit long-term use. Nutritional supplement in the form of Vitamin B6 may prove to be beneficial in a subset of patients.

## Materials and methods

The study included 25 individuals diagnosed with Tourette syndrome and 29 healthy individuals. The study was performed at the Department of Psychiatry and Neuroscience research laboratory at the Institute of Mental Health and Neurosciences (IMHANS). The study was approved by the Institutional Ethics Committee of the Government Medical College, Kozhikode. Written informed consent was obtained from the parents for participants under 18 years of age. Written informed consent was obtained from the control population as well.

### Inclusion criteria

25 Children between 5 and 18 years with Tourette syndrome as per DSM-5 criteria were included in the study. The Yale Global Tic Severity Scale was used to assess the severity and phenomenology of tics. ^8^The Children’s Yale-Brown Obsessive Compulsive Scale was utilized to evaluate OCD symptoms^.9^

Individuals with psychiatric disorders resulting from organic causes and substance abuse, intellectual disability, neurological, and neurodevelopmental disorders were excluded from the study.

An unrelated non-psychiatric healthy control group (age >20 years; n = 29) with no personal and family history of psychiatric disorders was recruited from the public.

### Sampling

10 mL of venous blood from the subjects was collected in an EDTA vacutainer for biochemical and molecular studies. Plasma and peripheral blood mononuclear cells (PBMC) were separated and stored at −80 °C deep freezer for further analysis. All plasma samples were protected from direct light exposure throughout processing and storage to prevent the photodegradation of the analyte. Plasma vitamin B6 levels were measured in 24 patients, as one blood sample was unavailable for analysis.

### Biochemical Analysis

The levels of vitamin B6 were measured from the individuals (Tourette syndrome and controls) in the plasma samples using ELISA following the manufacturer’s protocol. (GENLISA Human Vitamin B6 ELISA, KBH1539).

### Statistical Analysis

Quantitative variables were expressed as mean ± standard deviation as well as percentiles, as appropriate. The normality of data distribution was assessed using the Shapiro–Wilk test. Due to non-normal data distribution, comparisons between the two study groups were performed using the Mann–Whitney U test (two-tailed). A p-value < 0.05 was considered statistically significant.

## Results

There were 25 cases of Tourette’s syndrome, with a mean age of 11 years. 84% of the cases were males. 16% of the cases had ADHD or OCD as a comorbid diagnosis. 32% of the cases had a family history of Tourette syndrome, while 20% had a family history of OCD. Two children had a specific learning disorder (SLD).

The most common motor tics were eye blinking, shoulder shrugging, head jerking, and mouth movements. The common vocal tics were throat clearing, sniffing, uttering syllables, and breathing tics. Coprolalia was seen in four children.

Among the children with comorbid OCD, checking, washing, and symmetry were the predominant symptoms.

The mean age of females and males in this study was similar. Females did not have comorbid ADHD, OCD, or SLD, or a family history of Tourette’s. A single female had copropraxia and disinhibited behaviour. Complex tics were observed in 8 males and 1 female. These differences between females and males were not statistically significant.

Coprolalia was seen only in males. Among the males with coprolalia, two exhibited comorbid obsessive–compulsive disorder (OCD) and two presented with attention-deficit/hyperactivity disorder (ADHD). Two children had a family history of Tourette syndrome, while none had a family history of OCD. The median vitamin B6 levels in this subgroup did not differ significantly from those of other children with Tourette syndrome.

The control group had a mean age of 24 years and consisted of 22 males and 7 females. The median vitamin B6 level in the Tourette syndrome group was 25.01 ng/ml, compared with 36.33ng/ml in the control group. This difference was statistically significant (Mann–Whitney U = 225, p = 0.03). The rank-biserial correlation indicated a moderate effect size (r = 0.35)

**Table 1:**
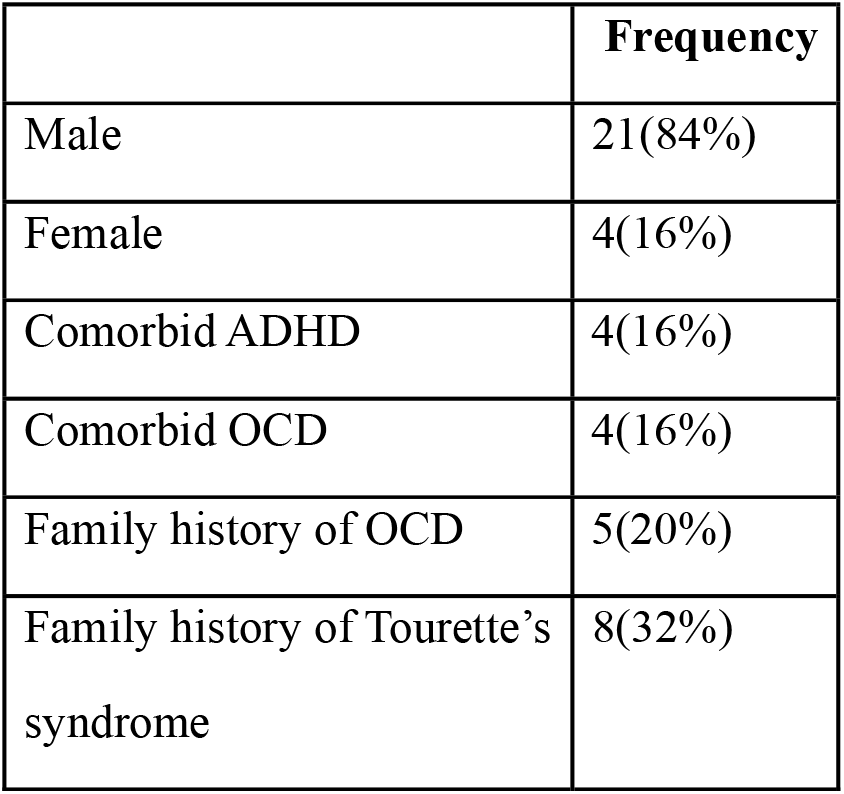
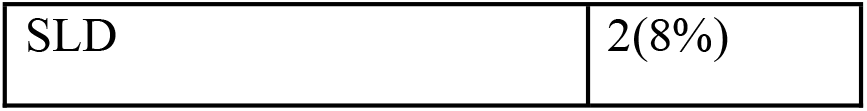
Clinical features.

**Table 2:**
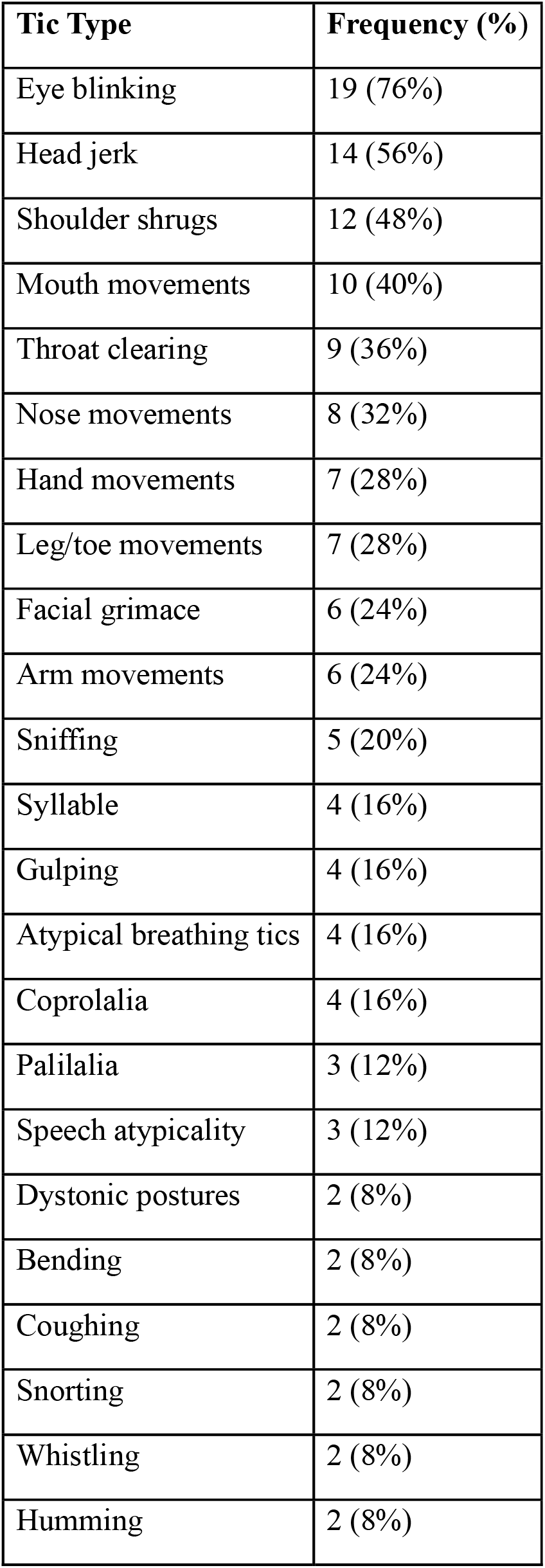
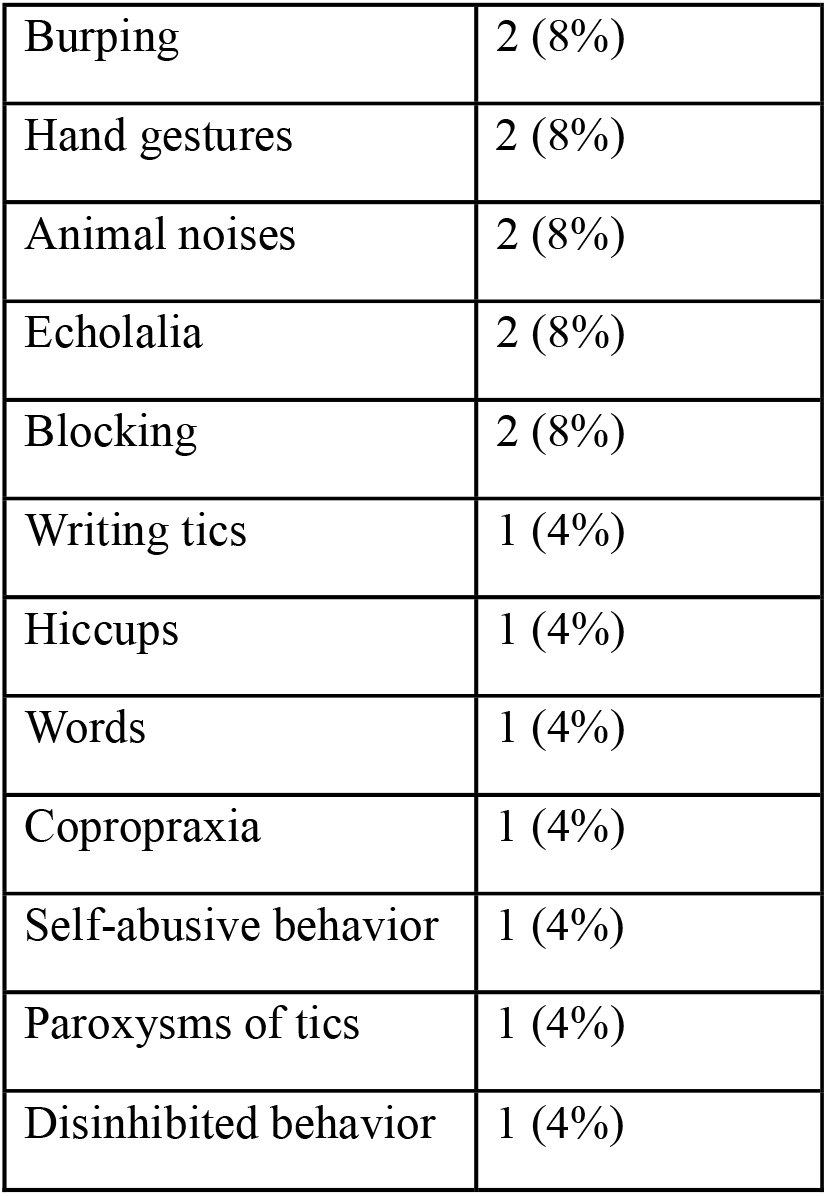
Phenomenology of tics.

**Table 3:** Plasma Vitamin B6 concentrations in TS cases and Controls.

|  | **Case** | **Control** |
| --- | --- | --- |
| N | 24 | 29 |
| Mean age (years) | 10.9 | 24.6 |
| Male | 20 | 7 |
| Female | 4 | 22 |
| Median B6 level in ng/ml (Inter Quartile Range) | 25.01(26.75) | 36.33(14.28) |
| Mann-Whitney U | 225 |  |
| P value | **0.03** |  |

## Discussion

This study assessed the clinical features of Tourette’s syndrome in children under 18 years and compared their vitamin B6 levels with those of normal controls.

Common motor tics in our study group were eye blinking, shoulder shrugging, and head jerking, while common phonic tics were throat clearing and sniffing. Complex tics, such as complex hand movements, were less frequent. Similar patterns have been reported in previous studies.^10^

Studies have shown that TS is one of the most heritable non-Mendelian neuropsychiatric disorders, with population-based heritability estimated to be 0.77. In twin studies, the concordance rate for TS is 53–56% in monozygotic twins, compared to 8% in dizygotic twins.^11^ Analyses of familial transmission patterns indicate that obsessive–compulsive disorder and TS share common genetic risk factors. ^2^ About one-third of patients had a family history of TS, while another one-fifth had a family history of OCD.

The study found that 16% of children with TS had comorbid OCD and ADHD, with these comorbidities occurring exclusively in males. Two males also displayed specific learning disorder. These findings align with global research, which shows that ADHD, OCD, learning difficulties, and disruptive behaviour are the common comorbidities in Tourette syndrome.^12^

OCD in TS showed predominance of sexual, religious, aggressive, or symmetry obsessions. ^13^ Symmetry obsessions were found in our group of patients in line with previous studies.

Tourette’s syndrome is more prevalent in males with earlier onset and a higher chance of ADHD and disruptive behaviour. In contrast, females are less likely to develop Tourette syndrome, have a later age of onset, and have more complex tics. They have less chance of comorbid ADHD but are more predisposed to comorbid OCD and mood symptoms. They also have a lower remission rate with increasing age. 84% of our cases were males with a mean age of 11 years.^14^ The mean age was similar for females and males in our study, differing from previous reports.

Complex tics were observed in 8 males and 1 female. This difference was not statistically significant. A study had found that other than ADHD, the frequency distribution and complexity of tics were similar in both genders. But in those below 18 years, earlier onset and complex tics were more common in males.^15^ Hyperandrogenism is postulated as the cause for the increased prevalence of TS in males. Sex specific neuroanatomic differences in the brain are less frequently reported in women with TS.^15^ Case reports show worsening of tics in men treated with androgenic steroids.^16^ Flutamide and finasteride, which are androgen blockers, showed improvement of tics in men. ^17,18^ All these points relate to the role of androgens in the causation of tics.

Coprolalia is a complex, socially inappropriate vocal tic where patients utter obscene words, while in copropraxia, patients show obscene gestures. Coprolalia and copropraxia are together called the copro phenomenon.^19^ The prevalence of coprolalia varies from 8% to 33% of patients with Tourette syndrome in various studies.^20^ Coprolalia is more than four times as common as copropraxia, and both are common in males. Copro phenomenon is associated with tic severity, aggression, self-injurious behavior, more brain dysfunction, and poor quality of life.^19^ In our study, coprolalia was observed in 16% of the patients-all being males. Two had a family history of TS. None had a family history of OCD. Two patients had comorbid ADHD, while two had OCD. Their mean vitamin B6, age of onset, did not differ from the remaining patients. Copropraxia was recorded in one female.

We found that the mean vitamin B6 level in the patients was less than that of the control, with a medium effect size. Whether this small amount is a causal factor in Tourette pathogenesis needs further evaluation. Vitamin B6 is involved in multiple biochemical pathways relevant to this disorder.

Tics occur due to increased dopamine in the striatal region.^2^ This increased dopamine may occur due to multiple pathways, with altered histaminergic metabolism being one. This is corroborated by studies showing altered signalling through H1 and H2 receptors in Tourette syndrome.^5^ Copy number variation analysis in a case-control study showed enrichment of genes within histamine receptors and GABA receptors.^21^Moreover, both H2 and H3 receptors are highly abundant in the human and rodent striatum and cortex—regions that have been implicated in the pathophysiology of Tourette syndrome.^22^In a subset of patients with a mutation in the histidine decarboxylase gene (HDC), reduced histamine levels increased striatal dopamine and resulted in tics.^23,24^ Vitamin B6 is a cofactor for the HDC enzyme.^24^ Other than a mutation in the enzyme, reduced B6 may impair the HDC enzyme activity, reduce histamine, and thus increase striatal dopamine. This provides a biologically plausible mechanism by which lower vitamin B6 levels could contribute to TS pathophysiology through the histamine pathway.

But impaired histamine metabolism is found only in a subset of TS. A genome-wide association study failed to identify any common polymorphisms with genome-wide statistical significance.^25^ In a post-mortem study transcriptome of the caudate and putamen of nine patients was assessed. They found a decrease in NOS (nitric oxide synthase) + interneurons, reduced voltage-gated potassium channel (KCNC1) in parvalbumin interneurons, and reduced GABA signalling genes.^26^ Vitamin B6 is a cofactor of glutamate decarboxylase, which synthesizes GABA^27^. Thus, other than histamine, reduced B6 may decrease GABA, thereby increasing the risk for TS.

In addition to its role in histamine and GABA synthesis, the immunoregulatory role of B6 may also play a role in TS pathogenesis. Genome-wide association studies have identified variants in FLT3, a key regulator of innate immune signalling, as being associated with TS.^28^Post-mortem transcriptomic analyses of the striatum have demonstrated an increased microglial inflammatory response ^26^. Vitamin B6 suppresses pro-inflammatory signalling pathways in activated microglia, including TLR4/NF-κB and TREM-1/DAP12–NLRP3–caspase-1–IL1β cascades, and reduces the production of reactive oxygen species, IL-6, and TNF-α.^29,30^ Together, these mechanisms suggest that vitamin B6 may mitigate genetically and transcriptionally driven neuroinflammatory processes implicated in TS. Supporting his hypothesis, a clinical trial in children with Tourette syndrome demonstrated a significant reduction in tics with vitamin B6 and magnesium supplements.^31^

This study is limited by a small sample size and differences in age and sex distribution between cases and controls, which may introduce residual confounding. In addition, plasma vitamin B6 levels may not accurately reflect central nervous system availability and the underlying neurobiological mechanisms relevant to Tourette syndrome.

Our study shows that TS in children is more common in males,with comorbid ADHD and OCD. Coprolalia, which causes significant distress to the patients, was found in a small subgroup. The vitamin B6 level in this group is lower, which may play a role in the disease pathogenesis. Future studies should incorporate a larger sample size with age and sex matched controls to reduce confounding. Measuring B6 level in CSF and identifying neuroinflammation using imaging would help in understanding the causal role of B6 in the pathophysiology of this disorder. Experiments using human microglial cell lines integrating transcriptomics and proteomics analysis will aid in identifying novel biomarkers. Finally, well-designed clinical trials with B6 supplementation will help in understanding its translational and therapeutic potential.

## Data Availability

All data produced in the present study are available upon reasonable request to the authors

## REFERENCE

1. Knight T, Steeves T, Day L, et al. Prevalence of tic disorders:a systematic review and meta-analysis. Pediatric Neurol 2012;47(2):77–90.

2. Du JC, Chiu TF, Lee KM, Wu HL, Yang YC, Hsu SY, Sun CS, Hwang B, Leckman JF. Tourette syndrome in children: an updated review. Pediatrics & Neonatology. 2010 Oct 1;51(5):255–64.

3. Baym CL, Corbett BA, Wright SB, Bunge SA. Neural correlates of tic severity and cognitive control in children with Tourette syndrome. Brain. 2008 Jan 1;131(1):165–79.

4. Lennington JB, Coppola G, Kataoka-Sasaki Y, Fernandez TV, Palejev D, Li Y, Huttner A, Pletikos M, Sestan N, Leckman JF, Vaccarino FM. Transcriptome analysis of the human striatum in Tourette syndrome. Biological psychiatry. 2016 Mar 1;79(5):372–82.

5. Rapanelli M, Pittenger C. Histamine and histamine receptors in Tourette syndrome and other neuropsychiatric conditions. Neuropharmacology. 2016 Jul 1;106:85–90.

6. Ercan-Sencicek AG, Stillman AA, Ghosh AK, Bilguvar K, O’Roak BJ, Mason CE, Abbott T, Gupta A, King RA, Pauls DL, Tischfield JA. L-histidine decarboxylase and Tourette’s syndrome. New England Journal of Medicine. 2010 May 20;362(20):1901–8.

7. Rapanelli M, Pittenger C. Histamine and histamine receptors in Tourette syndrome and other neuropsychiatric conditions. Neuropharmacology. 2016 Jul 1;106:85–90.

8. Leckman JF, Riddle MA, Hardin MT, Ort SI, Swartz KL, Stevenson JO, Cohen DJ. The Yale Global Tic Severity Scale: initial testing of a clinician-rated scale of tic severity. Journal of the American Academy of Child & Adolescent Psychiatry. 1989 Jul 1;28(4):566–73.

9. Scahill L, Riddle MA, McSwiggin-Hardin MA, Ort SI, King RA, Goodman WK, Cicchetti DO, Leckman JF. Children’s Yale-Brown obsessive compulsive scale: reliability and validity. Journal of the American Academy of Child & Adolescent Psychiatry. 1997 Jun 1;36(6):844–52.

10. Nilles C, Martino D, Fletcher J, Pringsheim T. Have we forgotten what tics are? A Re-exploration of tic phenomenology in youth with primary tics. Movement Disorders Clinical Practice. 2023 May;10(5):764–73

11. Liu S, Tian M, He F, Li J, Xie H, Liu W, Zhang Y, Zhang RU, Yi M, Che F, Ma X. Mutations in ASH1L confer susceptibility to Tourette syndrome. Molecular Psychiatry. 2020 Feb;25(2):476–90.

12. Robertson MM, Eapen V, Singer HS, Martino D, Scharf JM, Paschou P, Roessner V, Woods DW, Hariz M, Mathews CA, Črnčec R. Gilles de la Tourette syndrome. Nature reviews Disease primers. 2017 Feb 2;3(1):1–20.

13. Robertson MM. Tourette syndrome, associated conditions and the complexities of treatment. Brain. 2000 Mar 1;123(3):425–62.

14. Garris J, Quigg M. The female Tourette patient: sex differences in Tourette disorder. Neuroscience & Biobehavioral Reviews. 2021 Oct 1;129:261–8.

15. Baizabal-Carvallo JF, Jankovic J. Sex differences in patients with Tourette syndrome. CNS spectrums. 2023 Apr;28(2):205–11.

16. Bortolato M, Frau R, Godar SC, Mosher LJ, Paba S, Marrosu F, Devoto P. The implication of neuroactive steroids in Tourette’s syndrome pathogenesis: A role for 5α-reductase?. Journal of neuroendocrinology. 2013 Nov;25(11):1196–208.

17. Peterson BS, Zhang H, Anderson GM, Leckman JF. A double-blind, placebo-controlled, crossover trial of an antiandrogen in the treatment of Tourette’s syndrome. Journal of clinical psychopharmacology. 1998 Aug 1;18(4):324–31.

18. Muroni A, Paba S, Puligheddu M, Marrosu F, Bortolato M. A preliminary study of finasteride in Tourette syndrome. Movement disorders. 2011 Sep;26(11):2146–7

19. Kobierska M, Sitek M, Gocyła K, Janik P. Coprolalia and copropraxia in patients with Gilles de la Tourette syndrome. Neurologia i neurochirurgia polska. 2014 Jan 1;48(1):1–7.)

20. Goldenberg JN, Brown SB, Weiner WJ. Coprolalia in younger patients with Gilles de la Tourette syndrome. Movement Disorders: Official Journal of the Movement Disorder Society. 1994;9(6):622–5

21. Fernandez TV, Sanders SJ, Yurkiewicz IR, Ercan-Sencicek AG, Kim YS, Fishman DO, Raubeson MJ, Song Y, Yasuno K, Ho WS, Bilguvar K. Rare copy number variants in tourette syndrome disrupt genes in histaminergic pathways and overlap with autism. Biological psychiatry. 2012 Mar 1;71(5):392–402.

22. Deng H, Gao K, Jankovic J. The genetics of Tourette syndrome. Nature Reviews Neurology. 2012 Apr;8(4):203–13.

23. Ercan-Sencicek AG, Stillman AA, Ghosh AK, Bilguvar K, O’Roak BJ, Mason CE, Abbott T, Gupta A, King RA, Pauls DL, Tischfield JA. L-histidine decarboxylase and Tourette’s syndrome. New England Journal of Medicine. 2010 May 20;362(20):1901–8.

24. Baldan LC, Williams KA, Gallezot JD, Pogorelov V, Rapanelli M, Crowley M, Anderson GM, Loring E, Gorczyca R, Billingslea E, Wasylink S. Histidine decarboxylase deficiency causes tourette syndrome: parallel findings in humans and mice. Neuron. 2014 Jan 8;81(1):77–90.

25. Scharf JM, Yu D, Mathews CA, Neale BM, Stewart SE, Fagerness JA, Evans P, Gamazon E, Edlund CK, Service SK, Tikhomirov A. Genome-wide association study of Tourette’s syndrome. Molecular psychiatry. 2013 Jun;18(6):721–8.

26. Lennington JB, Coppola G, Kataoka-Sasaki Y, Fernandez TV, Palejev D, Li Y, Huttner A, Pletikos M, Sestan N, Leckman JF, Vaccarino FM. Transcriptome analysis of the human striatum in Tourette syndrome. Biological psychiatry. 2016 Mar 1;79(5):372–82.

27. Messripour M, Mesripour A. Effects of vitamin B6 on age associated changes of rat brain glutamate decarboxylase activity. Afr J Pharm Pharmacol. 2011 Mar 1;5(3):454–6.

28. Tsetsos F, Yu D, Sul JH, Huang AY, Illmann C, Osiecki L, Darrow SM, Hirschtritt ME, Greenberg E, Muller-Vahl KR, Stuhrmann M. Synaptic processes and immune-related pathways implicated in Tourette syndrome. Translational psychiatry. 2021 Jan 18;11(1):56.

29. Nguyen HD, Jo WH, Hong Minh Hoang N, Kim MS. The anti-inflammatory effects of vitamin B6 on neuroinflammation and neuronal damage caused by 1, 2-diacetylbenzene in BV2 microglial and sH-SY5Y cells. Immunopharmacology and Immunotoxicology. 2025 May 4;47(3):273–86.

30. Rakić M, Lunić T, Bekić M, Tomić S, Mitić K, Graovac S, Božić B, Nedeljković BB. Vitamin B complex suppresses neuroinflammation in activated microglia: in vitro and in silico approach combined with dynamical modeling. International Immunopharmacology. 2023 Aug 1;121:110525.

31. Garcia-Lopez R, Perea-Milla E, Garcia CR, Rivas-Ruiz F, Romero-Gonzalez J, Moreno JL, Faus V, Aguas GD, Diaz JC. New therapeutic approach to Tourette Syndrome in children based on a randomized placebo-controlled double-blind phase IV study of the effectiveness and safety of magnesium and vitamin B6. Trials. 2009 Mar 10;10(1):16.

